# Statistical Forecasting : Third Wave of COVID-19-With an Application to India

**DOI:** 10.1101/2021.12.20.21268150

**Authors:** Sabara Parshad Rajeshbhai, Subhra Sankar Dhar, Shalabh

## Abstract

The pandemic due to the SARS-CoV-2 virus impacted the entire world in different waves. An important question that arise after witnessing the first and second waves of COVID-19 is - Will the third wave also arrive and if yes, then when. Various types of methodologies are being used to explore the arrival of third wave. A statistical methodology based on the fitting of mixture of Gaussian distributions is explored in this paper and the aim is to forecast the third wave using the data on the first two waves of pandemic. Utilizing the data of different countries that are already facing the third wave, modelling of their daily cases data and predicting the impact and timeline for the third wave in India is attempted in this paper. The Gaussian mixture model based on algorithm for clustering is used to estimate the parameters.

## 1. Introduction

The COVID-19 pandemic impacted the entire world severely in an unprecedented way. The impact was so high that in many countries, the medical facilities were proven to be inadequate because the requirement was much beyond the expectations of Government. It had severe consequences in the form of spiralling cases, reduced supplies of essential treatments, and increased death toll. This also triggered the need of development of a new vaccine to control it and several vaccines, e.g., Covishield and Covaxin were developed and approved for emergency usage. The COVID-19 virus also started changing its mutation process and that resulted in a variety of symptoms in patients, degree of severity and the type of required medical attention. When the pandemic started, no one thought of having more than one waves. The first wave begun in early 2020 and it got subsided by later that year, it started again and the many countries witnessed the severe second wave of COVID-19. After witnessing the first and second wave, people started mentally preparing themselves to expect such more waves. This required scientific intervention from various fronts, e.g., medical science, biological science, virology, statistical science etc. The most important question that emerges is to explore the possibility of arrival of a third wave of COVID-19. This may require preparations on various fronts which will help in handling it in a better way.

Many people got interested to know the behaviour and pattern of the spread of coronavirus after the beginning of first wave of COVID-19 pandemic. People were more interested at that time to know when the pandemic will end believing that it will end. Nobody expected that there can be second and more waves also in the future. Again, as soon as the second wave started, people were interested once again in knowing its peak to know its impact and when it will finish. Another question cropped up by the time the second wave finished, people wanted to know can there be a third wave of the pandemic. By the time people could understand and realize, the third wave knocked at the doors of several countries. So a very important question is now knowing that there were two COVID-19 waves, when the third wave will arrive and finish.

This attracted many people working in different areas to use different types of tools to make forecasts. To answer the question to explore the possibility of arrival of third wave of COVID-19, people started using the mathematical models, biological models, statistical models etc. A major challenge in using the statistical tool is the availability of data. The statistical tools can do the forecasting on the basis of available data by creating a model and using it for the forecasting. A basic assumption behind the statistical forecasting is that the model which is obtained on the basis of the given set of data is assumed to continue to hold in the future and particularly, during the time period in which the forecasting is to be made. Another constraints in developing the statistical models is that they are derived from the data produced by the COVID-19 virus and many other factors. In a nutshell, the data is generated by the path which the nature adopts. The nature does not follow the statistical models. When the pandemic started, nobody knew how is the behaviour of the virus. Above all, they were many more factors that were impacting the propagation of virus. In simple words, we can say nobody knew how the nature is going to proceed further with this virus. So in the absence of any information to indicate the path of the nature, it was not easy to make any statistical forecast for the first wave or the second wave. As soon as some data started becoming available, people started using different types of statistical tools for modeling and to do related forecasting. We have also attempted in that direction, and we have used available data to make statistical forecasting about the arrival of third wave in India. Based on such a methodology, the forecasts for the other countries can also be made.

In the mean time, the vaccination also started and its effect started getting mixed up with the occurrence of COVID resulting in the less severity in the patients. Free vaccination campaign by the Government of India is indeed a good initiative but it will take some time to reach 100% efficacy. In many countries like the US, UK, Germany, Russia, majority of the people have been vaccinated but still they are currently facing the third wave. So, India and other countries have to build up the defences and be prepared for another wave so that it is not as devastating as the earlier ones.

This report is organized as follows: Section 2.1 describes the COVID-19 data source and try to visually get a sense of the trends and patterns present. The fitting of Gaussian mixture is explained in Section 2.2, and Section 2.3 discusses the prediction methodology. Section 3 contains the conclusion, and the report ends with a list of references.

## 2. Methodology and Analysis

Various statistical techniques can be used for forecasting. Among them, linear and nonlinear regression analysis, nonparametric regression, time series models etc. have been explored by various academicians. We attempt from a different perspective based on the nature of data.

### 2.1. Description of the Data set

We have considered the data from the first two waves of the COVID-19 pandemic from various countries available at https://ourworldindata.org/coronavirus, see [3]. The data on this website is updated daily and includes data on confirmed total cases, new cases, total and new deaths attributed to COVID-19, excess mortality as a percentage difference from previous years, hospital data such as number of COVID-19 patients in ICU, total number of COVID-19 patients in hospitals, and weekly new admissions of COVID-19 patients in hospitals, total tests conducted, daily new tests conducted, new and total single or fully vaccinations, and geographic data for all countries. The data is available in absolute values, per million of population as well as smoothed values. The “Our World in Data” relies on data from Johns Hopkins University, which is maintained by a team at its Center for Systems Science and Engineering (CSSE). It has been publishing updates on confirmed cases and deaths for all countries since January 22, 2020. The Johns Hopkins University updates its data multiple times each day. This data is sourced from governments, national and sub-national agencies across the world.

The data also has missing values which are not available on the website. For analysis purpose, we replaced any missing data point with the average of the previous and next day values.

Figure 1(a) shows India’s daily new cases per million, Figure 1(b) shows India’s daily new deaths attributed to COVID-19, Figure 1(c) shows daily new vaccination data for India, Figure 1(d) shows comparison between new tests and new cases for India, and Figure 1(e) shows the Stringency Index (a composite measure of the strictness of policy responses, 100 = strictest response)

**Figure 1:**
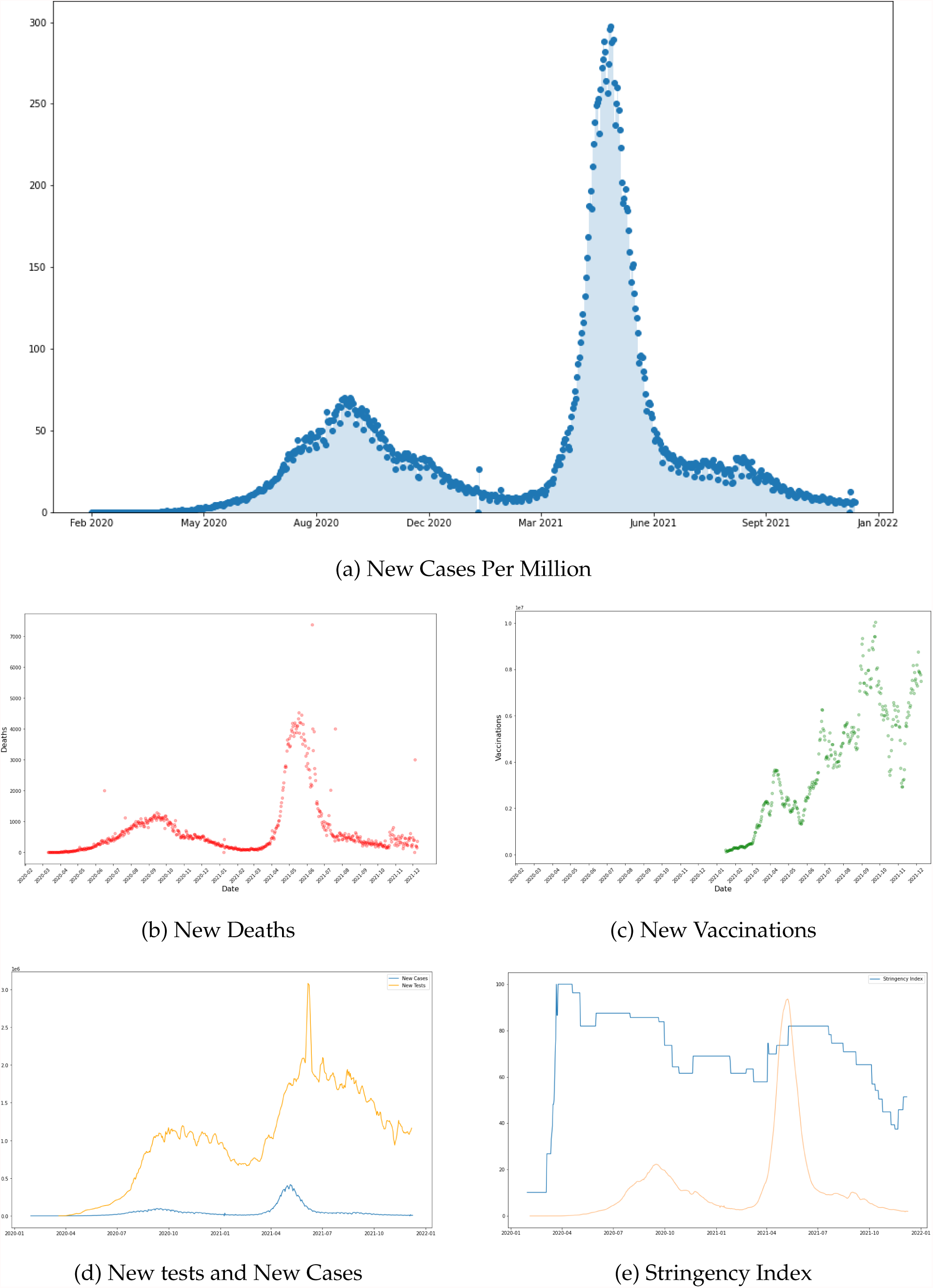
Visualizing the dataset

### 2.2. Fitting Gaussian Mixture on the data

Looking at the plot for daily new cases, it is clear that a linear model cannot map the data with a good fit and polynomial model may overfit the data of some countries while underfit others depending on the behaviour of COVID-19 waves in the different countries. While, probabilistic models of modelling the data seems like a good idea. Moreover, our belief is that the nature with respect to COVID-19 virus will follow more or less that the same path in different places depending on the local conditions. Looking at the path of the nature, a suitable distribution can be fitted and the forecasting can be made. The countries which are currently witnessing the third wave are selected. The reason of doing this is that our main objective is to the forecasting for India and India has already witnessed the two waves and expecting the third wave. Based on this assumption first we plotted the data of first and second waves in a two dimensional plot considering the number of cases with respect to time. The nature and structure of the curves for both the separate waves resembles with the Gaussian curves. When we try to combine the effects and data of both the first and second waves, then the the entire graph looks like a mixture of two normal distributions. So we plotted the related data for the number of cases with respect to time for different countries. Then we try to map the graph of India with those countries which have a similar pattern in the graphs. The graphs of different countries can be classified into several classes such that the graphs within the class resemble more to each other. We then plotted such a curve for India and tried to match it with a class to which it resembles the most. Then we mapped the data for the first two waves and fitted a mixture of Gaussian distributions. Using the best fitted distributions, we used the Mahalanobis distance between the means of individual distributions with an objective to forecast when the third wave will arrive in India. When we try to choose the curve which has got the best possible match and then we can believe that India is also going to follow us similar Curve.

After looking at the plots of daily cases, the graphs resemble with the Gaussian curves and can be assumed that the number of daily COVID cases follows a Gaussian distribution (a bell shaped curve). Gaussian distribution for a real-valued random variable can be given as: Let *X* ∈ ℝ follow a Gaussian distribution with mean *µ* and standard deviation *σ* i.e. *X* ∼ 𝒩(*µ,σ*),

For *x* ∈ ℝ

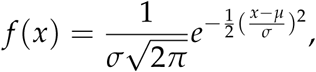

where *f* (.) is the probability density function. In case of India, the first wave seems shorter and spread out while the second curve is taller and squeezed together. So if the first wave follows a Gaussian distribution with mean *µ*_1_ and standard deviation *σ*_1_ and if the second wave follows a Gaussian distribution with mean *µ*_2_ and standard deviation *σ*_2_, then we can say that *µ*_1_ < *µ*_2_ and *σ*_1_ > *σ*_2_.

So, a Gaussian mixture of normal distributions can be a good model to fit on the COVID-19 data. The Gaussian mixture model (GMM) is well-known as an unsupervised learning algorithm for clustering, see [4]. Gaussian Mixture assumes that the data points come from multi-dimensional Gaussian distributions that could have different parameters. The idea is to find the parameters of the Gaussian distributions that best explain our data. This is known as generative modeling. We are assuming that these data are Normally distributed and we want to find parameters that maximize the likelihood of observing these data. In other words, considering each data as being generated by a mixture of Gaussian distributions, its probability can be calculated, see e.g., [1], [2].

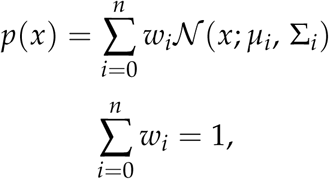

where *p*(*x*) is the probability density function, which is evaluated at *x*, of the random variable associated with number of deaths.

The first equation tells us that the density function evaluated at *x* is a linear combination of the *k* Gaussian distributions evaluated at x. We weight each Gaussian with *w*_*i*_, which represents the strength of that Gaussian. The second equation is a constraint on the weights: they all have to sum up to 1 to make the mixture distribution as a proper distribution. There are three different parameters that needs to be updated for the model: the weights for each Gaussian *w*_*i*_, the means of the Gaussian distributions *µ*_*i*_, and the variances of each Gaussian Σ_*i*_.

After solving these, it turns out that we can actually find converging values for the parameters! But the values of *w*_*i*_s have to be known beforehand. In other words, if it is known exactly which combination of Gaussian distributions at a particular point was taken from, then figuring out the means and variances is easy. But it is not the case in our analysis. There are two most important things in the Gaussian mixture model. One is to estimate the parameters for each Gaussian component within the Gaussian mixture and the other one is to determine which Gaussian component a data point belongs to. To achieve this, one can use the Expectation-Maximization algorithm. For this analysis, Gaussian Mixture object of scikit-learn package of python [4] was used.

Gaussian Mixture models work based on an algorithm called Expectation-Maximization, or EM. When given the number of clusters for a Gaussian Mixture model, the EM algorithm tries to figure out the parameters of these Gaussian distributions in two basic steps:

1. The E-step makes a guess of the parameters based on available data. Data points are assigned to a Gaussian cluster and probabilities are calculated that they belong to that cluster. The probability for a given Gaussian can be computed as: *w*_*i𝒩*_ (*x*; *µ*_*i*_, Σ_*i*_) and for normalization, dividing by 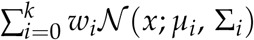
2. The M-step updates the cluster parameters based on the calculations from the E-step. The mean, variance and density are calculated for clusters based on the data points in the E step. It is to be noted that the GMM model of sklearn in Python uses covariance as the call parameter and it returns a matrix of variances of individual Gaussian distributions, see e.g., [2], [4]. To update a weight *w*_*i*_, we can simply sum up the probability that each point was generated by Gaussian *i* and divide by the total number of points. For a mean *µ*_*i*_, we can compute the mean of all points weighted by the probability of that point being generated by Gaussian *i*. For updating a variance Σ_*i*_, we can compute the variance of all points weighted by the probability of that point being generated by Gaussian *i*. Then perform these for each Gaussian *i*.

The two steps are repeated until convergence is reached.

### 2.3. Prediction

The first step of prediction is to check and shortlist which countries’ data of COVID-19 daily cases matches best with India. To check this, the daily new cases of all the countries are plotted against India to see which countries have their first two waves behaving in a similar manner with India. In the case of India, there are two distinct waves where first wave is more spread out (with greater standard deviation) while the second wave is more squeezed together (lesser standard deviation). Also, considering a mixture of Gaussian distribution, the weight factor for the first wave looks lesser than the weight factor for the second wave.

Fitting a mixture of Gaussian distribution to the data gives the mean, standard deviation and weight factors for each individual Gaussians. To predict the timeline of the third wave in India, the Mahalanobis distance between the peak case dates (mean) for the training country dataset are used to predict the peak case date of the third wave. The difference between the overall duration of the waves (standard deviations) are used to predict the standard deviation for India’s third wave. Taking the base values of the weight factor, the M-step of the EM algorithm is used to predict the weight factor of the third wave.

From Figure 2(a) and Figure 2(b), it is clear that South Africa and Zimbabwe’s curve (orange) have distinct first two peaks, similar to India (blue). Whereas, in Figure 2(c) and Figure 2(d), Netherlands and Norway’s curve (orange) do not have distinct peaks for first two waves that are similar India (blue).

**Figure 2:**
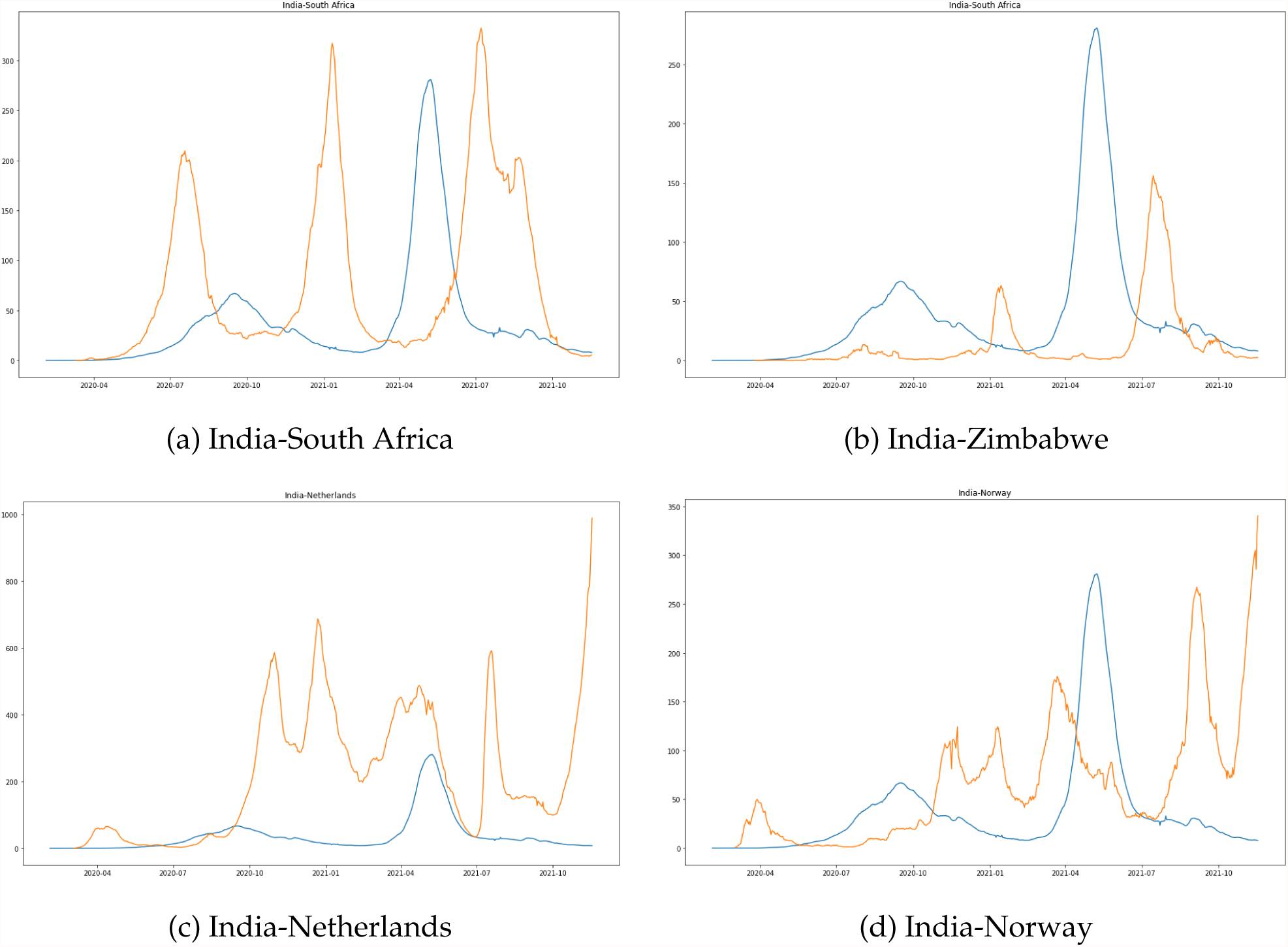
Matching daily cases data of different countries with India

After plotting the daily cases per million of all other countries and matching the graphs with India, top 10 countries which matched the best are chosen as a training dataset. The top 10 countries are: USA, UK, Germany, France, South Africa, Russia, Israel, Spain, Zambia and Zimbabwe. Zambia and Zimbabwe are the closest matching countries for which the daily cases data follow very similar pattern to India’s daily cases data.

Figure 3 shows the curves of the top 10 countries’ new cases per million population data and matches with India’s data. India’s data is shown in blue while other countries are shown as orange.

**Figure 3:**
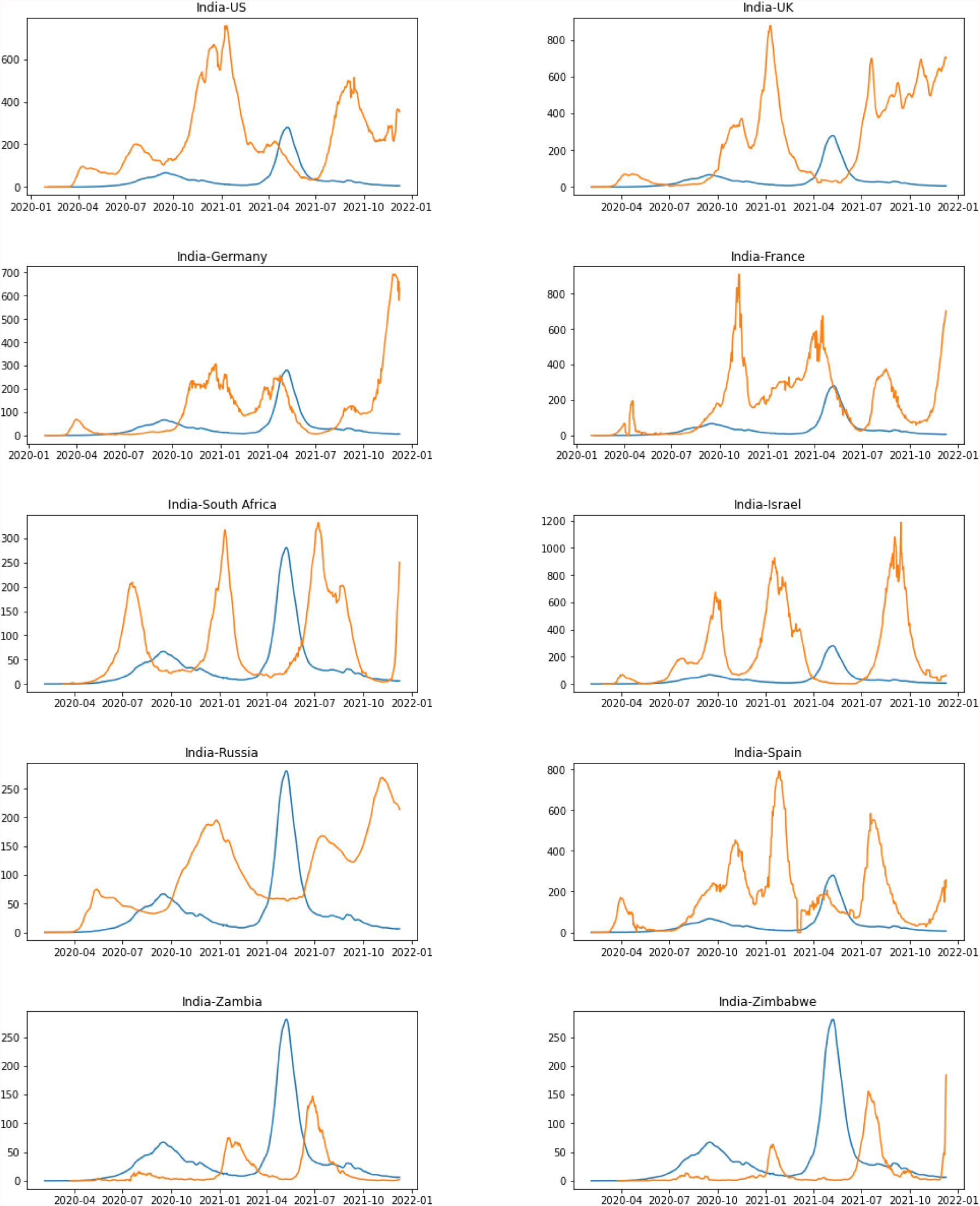
Top 10 countries that match best with India

**Figure 4:**
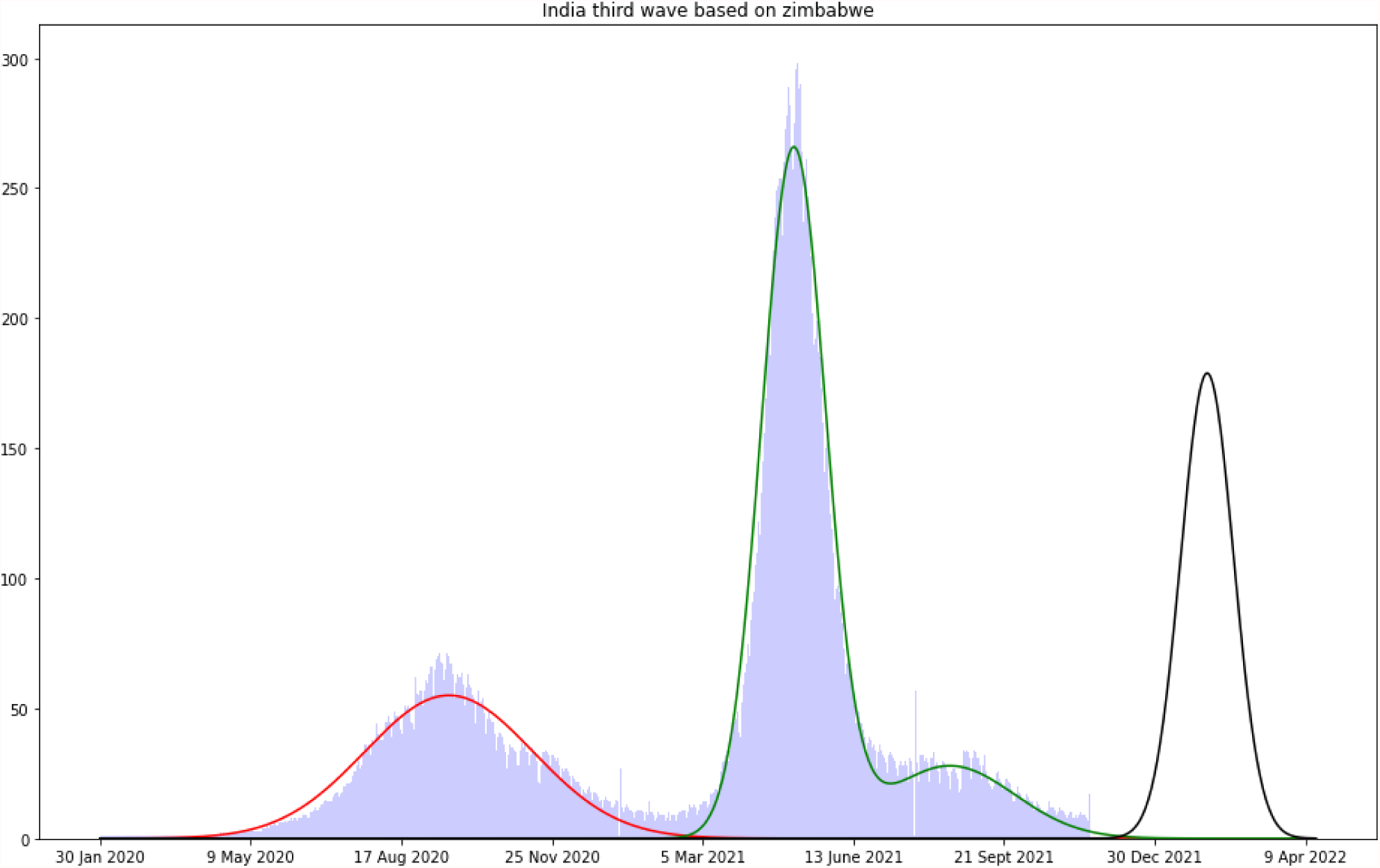
Prediction of India’s third wave based on Zimbabwe

After fitting a mixture of Gaussian Distribution to the dataset, we obtain the values of the mean, standard deviation and individual weights for each individual Gaussian distributions that represent the given data.

For training the models, chronologically numbered format for dates instead of date-time formats is used. Using the Gaussian Mixture model for fitting India’s data, we get a mixture of two Normal distributions as expected. *µ*_1_ ≈ 232, *σ*_1_ ≈ 56 and *µ*_2_ ≈ 460, *σ*_2_ ≈ 21. Using the same approach to model the data for other countries, we get three sets of parameters for the countries denoting the three waves of COVID-19 cases.

Using the prediction approach mentioned above, the parameters for the third wave of COVID-19 in India are predicted. Taking Zimbabwe as training set, the parameters of the third wave are:

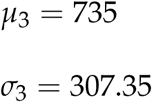

This suggests that the cases reach the peak value after 735 days from our initial observation date which is 30 January 2020. So, the cases start rising around 15 December 2021 and the peak of the third wave will occur on Thursday, 3 February 2022.

## 3. Conclusion

After facing severe consequences of the second wave of COVID-19, India has to be prepared for a third wave even if the chances of a third wave hitting the country is quite low. Assuming that a third wave of COVID-19 may hit the country, and following the trends happening around the world, using Gaussian Mixture model to fit the COVID-19 data and make predictions, this project report forecasts India’s third wave of COVID-19 to start around mid December 2021 and the cases to peak in the beginning of February 2022.

### 3.1. Future Work

While the timeline can be estimated with some confidence using this approach, the exact peak number of cases (weight factor) cannot be estimated accurately as it involves the vaccination data of the population. So, modeling of the total vaccination data can be done using a Linear Quantile Regression Model for Zimbabwe and India and it can be used for accurately estimating the peak number of cases. Also, since this report considers only one country Zimbabwe as our training data set, it can be extended it to other countries as well and an average of the parameters can be used for the prediction of third wave of COVID-19 in India.

## Data Availability

https://ourworldindata.org/coronavirus

## ^1^Acknowledgement

The authors gratefully acknowledges the support from the MATRICS project from Science and Engineering Research Board (SERB), Department of Science and Technology, Government of India.

## References

[1] Mohit Deshpande. Clustering with gaussian mixture models. 2020. URL https://pythonmachinelearning.pro/clustering-with-gaussian-mixture-models.

[2] Vivienne DiFrancesco. Gaussian mixture models for clustering. 2021. URL https://towardsdatascience.com/gaussian-mixture-models-for-clustering-3f62d0da675.

[3] Lucas Rodés-Guirao Cameron Appel Charlie Giattino Esteban Ortiz-Ospina Joe Hasell Bobbie Macdonald Diana Beltekian Hannah Ritchie, Edouard Mathieu and Max Roser. Coronavirus pandemic (covid-19). Our World in Data, 2020. URL https://ourworldindata.org/coronavirus.

[4] F. Pedregosa, G. Varoquaux, A. Gramfort, V. Michel, B. Thirion, O. Grisel, M. Blondel, P. Prettenhofer, R. Weiss, V. Dubourg, J. Vanderplas, A. Passos, D. Cournapeau, M. Brucher, M. Perrot, and E. Duchesnay. Scikit-learn: Machine learning in Python. Journal of Machine Learning Research, 12:2825–2830, 2011. URL https://scikit-learn.org/stable/index.html.

